# Perceived Barriers and Facilitators of Using Synchronous Telerehabilitation of Physical and Occupational Therapy in Musculoskeletal Disorders: A Scoping Review

**DOI:** 10.1101/2022.07.21.22277858

**Authors:** Lydia Tao, Andrea Carboni-Jiménez, Kimberly Turner, Nora Østbø, Kylene Aguila, Jill Boruff, Marie-Eve Carrier, Ankur Krishnan, Christiane Azar, Andréanne Guindon, Natacha Viens, Sara Ahmed, Brett D. Thombs, Linda Kwakkenbos

## Abstract

**Purpose:** Physical and occupational therapy interventions are increasingly delivered through videoconferencing to overcome barriers related to face-to-face delivery. The objective of this scoping review was to identify barriers and facilitators of using synchronous telerehabilitation to deliver these interventions for musculoskeletal disorders.

**Materials and Methods:** MEDLINE, EMBASE, PsycInfo, CINAHL, Cochrane Library, and ProQuest Dissertations and Theses databases were searched in May 2020. Qualitative and quantitative studies in any language that described barriers and facilitators of using synchronous videoconferencing for physical or occupational interventions or assessments for individuals with musculoskeletal diseases were eligible.

**Results:** Twenty-three publications were included that reported 59 facilitators and 41 barriers to using telerehabilitation. All included studies (100%) reported on facilitators, and 20 (87%) studies also reported on barriers. Most commonly reported facilitators included convenience and accessibility of services, audio and visual quality, and financial and time savings. Most commonly reported barriers included technological issues, privacy concerns, impersonal connection, and difficulty establishing rapport between patients and healthcare professionals.

**Conclusions:** Factors including quality and user-friendliness may facilitate the delivery of physical or occupational therapy interventions or assessments for musculoskeletal diseases using telerehabilitation. Strategies to address key barriers should be considered when developing and implementing such interventions or assessments.

**Implications for rehabilitation:**

- Videoconferencing with a healthcare professional can be an effective way to deliver patient-centered physical or occupational therapy telerehabilitation interventions.
- Strategies to combat barriers to using telerehabilitation may include using a stable, high-quality videoconferencing platform, enhancing self-efficacy to using videoconferencing amongst patients and health care providers, and addressing concerns related to privacy.
- During the current COVID-19 pandemic, the present study provides insight into the successful development and delivery of physical or occupational telerehabilitation interventions for at-risk populations.

## Introduction

Rehabilitation interventions play a crucial role in limiting disability and improving health-related quality of life (HRQL) in chronic diseases. In many chronic diseases, exercise, rehabilitation, and self-management programs are effective complements to basic medical care.^2-4^ People with rare diseases, however, often have difficulty accessing appropriate services.^5^ Rare diseases are chronic, disabling medical conditions that affect fewer than 1 in 2,000 people.^6,7^ Approximately 70% of rare diseases have fewer than 1,000 cases worldwide.^8^ Nonetheless, overall, 6-8% of the world’s population may have a rare disease.^6,7^ The burden and impact on daily function and HRQL of most rare diseases is high.^9,10^

Most rare diseases have no therapy that cures or modifies the disease itself.^9^ Rehabilitation interventions, such as physical and occupational therapy could potentially help rare disease patients achieve better physical function and HRQL, but the small number of patients with any single rare disease^7^ is a barrier to the development and testing of effective disease management and rehabilitation tools. Country-specific national rare disease plans emphasize the need to develop, test, and disseminate programs to improve the ability of people to manage and cope with rare diseases.^11-14^ No national plans, however, have proposed a structure for how to do this feasibly and cost-effectively, which is an important consideration in the context of small patient numbers in any clinical setting or country, as well as the limited resources available. Thus, finding a way to effectively develop, test, and deliver patient-centered interventions for patients with rare diseases is an important, but unsolved problem.

Many rare diseases have musculoskeletal implications.^15,16^ Systemic sclerosis (SSc), for example, is a rare autoimmune connective tissue disease characterized by significant musculoskeletal involvement.^17,18^ Musculoskeletal rehabilitation and physical and occupational therapy are recommended for the management of musculoskeletal impairment in SSc.^19^ Rehabilitation activities that have been recommended, however, are typically based on small randomized controlled trials or case reports. They include range of motion exercises, including hand and orofacial exercises; connective tissue massages; joint manipulation; splinting; and heat or paraffin wax baths.^19-21^ A trial of 220 patients with SSc found that a 4-week general physical therapy program significantly reduced disability 1 month post-randomization, although there was no effect on disability at 12-month follow-up.^22^ A recent study of 1,627 SSc patients included in the international Scleroderma Patient-centered Intervention Network (SPIN) Cohort, however, found that fewer than 25% of participants had used physical or occupational therapy services in the 3 months prior to study enrollment.^23^

Telerehabilitation interventions delivered via videoconferencing with a healthcare professional are increasingly common and effective for addressing a range of healthcare problems.^24-27^ Using telerehabilitation interventions to address disability and functional limitations is a promising approach to providing disease-specific physical and occupational therapy interventions and has been proven to be reliable and effective for assessment, treatment, and postoperative follow-ups for musculoskeletal diseases.^28^ However, knowledge is needed on facilitators and barriers to successful implementation. To obtain the necessary knowledge to develop, test and effectively disseminate physical or occupational therapy interventions or assessment protocols for people with SSc and other common or rare musculoskeletal conditions, we conducted a scoping review to identify barriers and facilitators of using telerehabilitation methods for rehabilitation in musculoskeletal diseases.

## Methods

A scoping review is a “form of knowledge synthesis that addresses an exploratory research question aimed at mapping key concepts, types of evidence, and gaps in research related to a defined area or field by systematically searching, selecting, and synthesizing existing knowledge.”^29^ A scoping review is rigorous like a systematic review; however, unlike a systematic review, it addresses broader topics and charts all available evidence, regardless of study design or quality.^30^ The scoping review was conducted following the approach described by Arksey and O’Malley,^30^ which has since been refined by others.^29,31^ Steps in the process include (1) identifying the research question, (2) identifying eligible studies, (3) study selection, (4) charting the data, and (5) collating, summarizing, and reporting results.^29,31^

### Identifying the Research Question

To guide this scoping review, we defined the following research question: What are barriers and facilitators of using telerehabilitation methods to deliver physical or occupational therapy interventions or assessments for musculoskeletal diseases?

### Eligible Studies

Eligible publications were required to describe barriers or facilitators of using telerehabilitation methods in physical or occupational therapy for individuals with a musculoskeletal disease. Only articles about diseases of the musculoskeletal system and connective tissue, defined per the International Statistical Classification of Diseases and Related Health Problems, 10th revision (ICD-10; M00-M99) were included.^32^

For the purpose of this study, telerehabilitation interventions, sometimes described as telehealth or e-health interventions, were defined as delivery of care from a distance to support, educate, inform and connect health care professionals and the people they serve through the use of information communications technologies. Eligible intervention delivery and assessment methods included online video conferencing, online interventions, mobile phone apps (mHealth), and the use of remotely monitored rehabilitation devices to deliver online rehabilitation interventions. For this review, telerehabilitation methods had to involve a component of direct, synchronous clinician and patient interaction via telerehabilitation methods, which may or may not have also included self-guided online material. Interventions that involved telephone interactions only were excluded.

Eligible occupational therapy interventions included interventions described as an occupational therapy intervention and interventions delivered by an occupational therapist or under the supervision of an occupational therapist. Occupational therapy interventions focus on activities of daily living as well as instrumental activities of daily living. Eligible occupational therapy assessments were those conducted by an occupational therapist for the purpose of designing, adapting, or evaluating an occupational therapy intervention. Eligible physical therapy interventions included interventions described as a physical therapy intervention and interventions delivered by a physical therapist or under the supervision of a physical therapist. Eligible physical therapy assessments were those conducted by a physical therapist for the purpose of designing, adapting, or evaluating a physical therapy intervention.

Consistent with standard scoping review methodology, we did not include any study design restrictions.^29,31^

### Identifying Relevant Studies

To identify potentially relevant publications describing barriers and facilitators to using telerehabilitation methods to deliver physical or occupational therapy for musculoskeletal conditions we searched MEDLINE (Ovid), EMBASE (Ovid), PsycInfo (Ovid), CINAHL (EBSCO), Cochrane Library, and ProQuest Dissertations and Theses databases from inception until May 27, 2020 with no language restrictions. A librarian with expertise in systematic and scoping review searching developed the search strategy and performed the search. MEDLINE strategies for the search were developed with input from the project team and were peer reviewed using the Peer Review of the Electronic Search Strategy standard.^33^ The MEDLINE strategy was then adapted for other databases, tailoring vocabulary and syntax to allow for optimal electronic searching. The search strategy can be found in Appendix 1.

### Study Selection

The results of the searches were downloaded into the citation management database RefWorks,^34^ and duplicate references were identified and removed. Following this, references were transferred into the systematic review software DistillerSR.^35^ A coding manual based on eligibility criteria was developed and pilot-testing of the coding manual was performed prior to the study’s inception. The coding manuals are shown in Appendix 2.

The eligibility of each publication was assessed through a two-stage process. First, two investigators independently reviewed the titles and abstracts of all citations identified through the search strategy using a liberal accelerated method^36^ to screen titles and abstracts, meaning that articles deemed eligible by one of the reviewers were included in full-text review, and only excluded articles were screened by a second reviewer. Since title and abstract screening was done randomly and concurrently, reviewers did not know if the other reviewer had excluded the reference or not. In the second stage, two investigators independently conducted a full-text review of all articles. Disagreements after full-text review were resolved by consensus, with a third investigator consulted, as necessary.

### Charting the Data, and Collating, Summarizing, and Reporting Results

A descriptive analytical approach was used to chart and summarize the data from included publications. For each publication, we extracted: (1) authors; (2) publication year; (3) country; (4) study design; (5) data collection methods; (6) number of participants; (7) musculoskeletal condition(s); (8) brief description of participants; (9) type of telerehabilitation intervention; (10) features or components of the telerehabilitation intervention and underlying behaviour change mechanism; (11) summary of outcomes; (12) facilitators; and (13) barriers.

The Consolidated Framework for Implementation Research (CFIR) was used as a guide for categorizing barriers and facilitators.^37^ The CFIR is a commonly used conceptual framework that lists 39 constructs in five domains thought to influence implementation of interventions. Two investigators independently extracted data from included publications and entered it into a standardized Excel spreadsheet. Afterwards, they categorized each barrier and facilitator within the CFIR domains (*Intervention Characteristics, Outer Setting, Inner Setting, Characteristics of Individuals*, and *Process*) and the corresponding constructs under each domain.^*37*^ *Intervention Characteristics* refers to the key intervention features that may influence the success of implementation (e.g., design quality, cost, complexity). *Outer Setting* describes the external influences on intervention implementation (e.g., patient needs and resources, external policy, and incentives). *Inner Setting*, on the other hand, refers to the implementation climate, culture, and other features such as leadership engagement inside of the implementing organization. *Characteristics of Individuals* relate to, for example, knowledge and belief about the intervention, self-efficacy of individuals, and other personal attributes. *Process* refers to different implementation phases, including planning, engaging, executing, reflecting, and evaluating, as well as strategies during which that might affect the implementation. Any disagreements after coding and categorizing were discussed and resolved, with a third investigator consulted as necessary.

## Results

The database searches yielded 1728 unique citations. Of these, 1464 articles were excluded after the title and abstract review, leaving 264 publications for full-text review. Of these, 23 publications were included in the scoping review (see figure 1).

**Figure 1:**
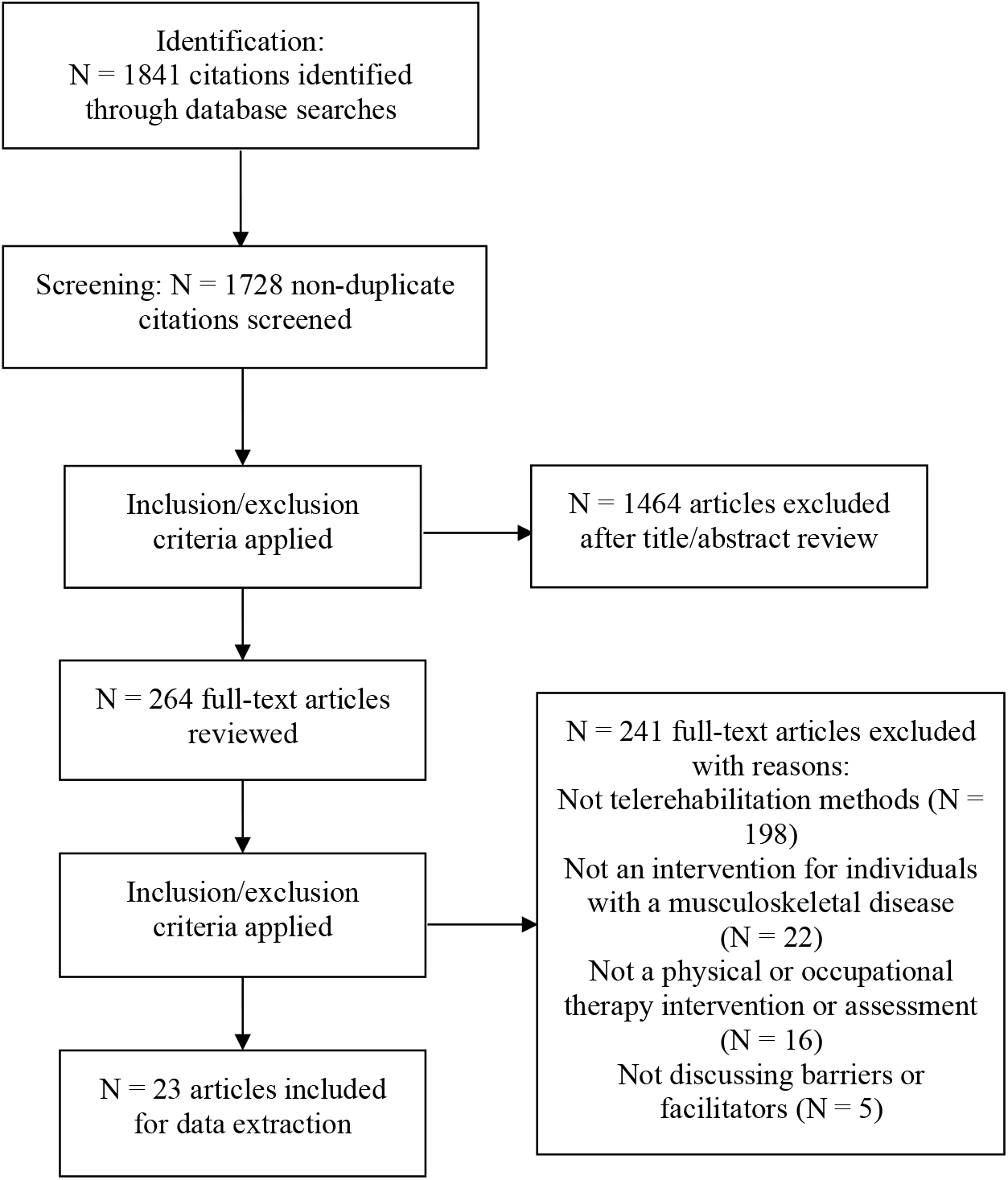
Flow diagram of the study identification and screening.

### Description of Included Studies

Of the 23 included studies, 21 (91%) were primary research studies,^38-43,45-52,54-60^ and two (9%) were systematic reviews.^44,53^ Of the 21 primary studies, there were eight randomized controlled trials (RCTs) (38%),^39,43,47,50,51,54,57,58^ five mixed-methods studies (24%),^42,48,52,55,60^ five observational studies (24%),^38,46,49,56,59^ and three qualitative studies (14%).^40,41,45^ Musculoskeletal conditions of study participants in the 23 studies included osteoarthritis (*n* = 8, 35%),^39,43,45,46,49-51,53^ lower back pain (*n* = 3, 13%),^41,52,59^ ankle pain and dysfunction (*n* = 2, 9%),^41,56^ knee pain (*n* = 2, 9%),^41,60^ shoulder pain (*n* = 2, 9%),^41,57^ and other or general musculoskeletal conditions (*n* = 9, 39%).^38,40,42,44,47,48,54,55,58^. One study^41^ included participants with multiple musculoskeletal conditions. Fifteen studies (65%) were conducted with participants from Australia,^38-41,45,46,48,49,51,53-57,59^ five (22%) were from Canada,^42-44,50,58^ and there was one each from South Korea (4%),^47^ Spain (4%),^52^ China (4%).^60^ All publications were from 2005 or later.

The two systematic reviews evaluated the effect of physiotherapy exercise and activity interventions delivered via videoconferencing among included studies.^44,53^ Of the 21 primary studies, 17 studies described physical therapy interventions including 1 physical therapy consultation,^38^ 7 assessments or evaluations,^48,52,55-59^ and 9 treatments;^39,43,45,47,49-51,54,60^ One study described a multidisciplinary telerehabilitation program involving both physical and occupational therapy consultations.^42^ One study involved pre-admission consultations between patients and occupational therapists.^46^ In two studies, features or components of the telerehabilitation intervention were not reported.^40,41^ Characteristics of included publications are summarized in table 1.

**Table 1.** Characteristics of included studies (N = 23)

| First author, year | Country | Design | Data collection method(s) | Musculoskeletal condition(s) | Features of the telerehabilitation intervention | Summary of outcomes | Participants |
| --- | --- | --- | --- | --- | --- | --- | --- |
| Beard, 2017 <sup>38</sup> | Australia | Observational study | Data collection forms and phone follow-ups | Spinal disorders | Patients in the telehealth group received weekly telehealth consultations (3 hours) via videoconference over 5 months. During the videoconference, the Lead Physiotherapist clarified clinical features and discussed assessment and imaging findings with the patient and local physiotherapist. Then, a consensus view regarding the provisional diagnosis and management recommendations were provided to the patient with any remaining questions addressed. | Outcomes included analysis of process, service activity (wait-list times, diagnosis, attendance rates, discharge rates, imaging and intervention rates), clinical actions, safety and costs. | 41 patients, including 19 enrolled into the Spinal Assessment Clinic Outreach and 22 into the Spinal Assessment Clinic Telehealth, referred by a general practitioner for assessment of a spinal disorder from the Royal Adelaide Hospital spinal service. |
| Bennell, 2017 <sup>39</sup> | Australia | Randomized controlled trial (RCT) | Not specified | Knee osteoarthritis | Online educational material and 7 Skype sessions (45-minutes) with a physiotherapist over 12 weeks and an online pain-coping skills program. | Primary outcomes were pain during walking and physical function at 3 months. Secondary outcomes were knee pain, quality of life, global change (overall, pain, and functional status), arthritis self-efficacy, coping, and pain catastrophizing. | 74 patients in control group (mean age = 61.5, SD = 7.6) and 74 patients in intervention group (mean age = 60.8, SD = 6.5). |
| Cottrell, 2017a <sup>40</sup> | Australia | Qualitative study | Interview | Chronic musculoskeletal conditions | N/A | (1) current perceived barriers to patients' accessing the Neurosurgical & Orthopaedic Physiotherapy Screening Clinic and Multidisciplinary Service; (2) whether telerehabilitation could address these barriers; (3) potential barriers and facilitators to successfully implement telerehabilitation. | 26 health care professionals: Physiotherapy (N=15), Occupational therapy (N=2), Nutrition & Dietetics (N=4), Psychology (N=4), Pharmacy (N=1). |
| Cottrell, 2017b <sup>41</sup> | Australia | Qualitative study | Questionnaire | Low back pain, neck/thoracic pain, shoulder pain, knee | N/A | (1) current barriers to attendance to medical appointments; (2) satisfaction with current management within the | 84 patients (45 females and 39 males) receiving services from the Neurosurgical and |
|  |  |  |  | pain, hip pain, other (ankle/foot and hand) |  | Neurosurgical and Orthopaedic Physiotherapy Screening Clinic and Multidisciplinary Service; (3) technology access and literacy; and (4) attitudes and preferences towards telehealth. | Orthopaedic Physiotherapy Screening Clinic and Multidisciplinary Service. |
| Dickson, 2008<sup>42</sup> | Canada | Mixed-methods study | Questionnaire and interview | Osteoporosis | The telehealth component of the intervention included mock consultations between healthcare professionals and pseudo patients, as well as the actual consultations with real patients. Patients received an approximately 2-hour consultation with the healthcare team (patients met each professional in the healthcare team individually), via a set-top Tandberg 880 videoconferencing system with a 27-inch television in the NORTH network studio. | Outcomes were the feasibility of delivering a multidisciplinary model of care through telehealth and ways to improve access to specialist care for osteoporosis investigation and management. | 20 patients (18 women and 2 men; mean age = 56.5) referred to the osteoporosis telehealth program by family physicians. |
| Doiron-Cadrin, 2020<sup>43</sup> | Canada | Randomized controlled trial (RCT) | Questionnaire | Hip or knee osteoarthritis | Patients received prehabilitation sessions over 12 weeks under the supervision of a physiotherapist through telecommunication applications (two sessions per week). | Primary outcome was the Lower Extremity Functional Scale (LEFS). Secondary outcomes were feasibility, patients' acceptance and compliance to the program, the Western Ontario and McMaster Universities Osteoarthritis Index (WOMAC), the Short Form Health Survey (SF-36), the Self-Pace Walk, the Stair Test, the Timed Up and Go, and a Global Rating of Change scale. | 34 French-speaking adults (9 males and 25 females) with severe hip or knee osteoarthritis from wait lists for a hip or knee total joint arthroplasty in Quebec, Canada. 12 were allocated to the tele-prehabilitation group, 11 to the in-person group and 11 to the control group. |
| Grona, 2017<sup>44</sup> | Canada | Systematic Review | Review of primary studies | Musculoskeletal conditions (general) | Physical therapy assessment or treatment. | (1) the reliability and validity of videoconferencing for physical therapy management of musculoskeletal conditions and (2) the health, system, and process outcomes when using secure videoconferencing for physical therapy management of musculoskeletal conditions | Adults 18–80 with chronic musculoskeletal disorders (>3 months duration) from 17 studies included in systematic review. |
| Hinman, 2017<sup>45</sup> | Australia | Qualitative study | Interview | Knee osteoarthritis | Participants received 7 internet-based Skype-delivered physical therapy sessions for 3 months, the main purpose being to prescribe an individualized home-based | Outcomes were patients' and therapists' perspectives and experiences related to quality of healthcare (i.e., structure, process, and outcomes). | 12 patients (6 men and 6 women; mean age = 62) who had mild to moderate OA symptoms, and 42% (n = 5) lived in rural areas of Australia. |
|  |  |  |  |  | strengthening program to be undertaken 3 times per week. In the first session, the therapist selected 5–6 suitable exercises from the study protocol and demonstrated them, then the participant performed the exercise while the physical therapist watched. At subsequent sessions, exercises were reviewed and progressed. |  | 8 physical therapists (4 men and 4 women) who had an average of 15 years of clinical experience. |
| Hoffmann, 2008 <sup>46</sup> | Australia | Observational study | Questionnaire and interview | Osteoarthritis | A face-to-face occupational therapist brought the equipment to the patient's home, set it up, established a dial up internet connection, and explain how to use the equipment to them. Patients were then invited to operate the equipment throughout the online home visit performed by an online therapist and the face-to-face therapist. During the online visit, the online therapist directed the face-to-face therapist to take video pictures of various areas of the house, asked patients to demonstrate certain transfers, and took furniture height measurements, and also asked patients questions. Both therapists completed the same home environment questionnaire after home visit completed. | Primary outcome was agreement between remote and face to face occupational therapist ratings on the Home Environment Questionnaire. Secondary outcome was how the participants felt about using the equipment. | 40 patients (14 males, 26 females; mean age = 68) who were scheduled to undergo either total hip or total knee replacement participated in the study. |
| Hong, 2017 <sup>47</sup> | South Korea | Randomized controlled trial (RCT) | Not specified | Sarcopenia | The tele-exercise group performed supervised resistance exercise at home for 20–40 min a day three times per week for 12 weeks. The remote instructor provided one-on-one instruction to each participant during the intervention via the video conferencing software (Skype™). The control group maintained their lifestyles without any special intervention. | Primary outcomes were body composition and functional fitness measured at baseline and endpoints (post 12-week intervention). | 11 patients (5 males and 6 females; mean age = 82.2) from the treatment group and 12 patients (5 males and 7 females; mean age = 81.5) in control group. |
| Lade, 2012 <sup>48</sup> | Australia | Mixed-methods study | Observation and questionnaire | Elbow injury or pain | Remote physical examinations guided by a remote examiner using a commercial telerehabilitation system. | Primary outcomes were the validity and reliability of a telerehabilitation examination and diagnosis of elbow musculoskeletal disorders. Secondary outcomes were physical examination test results, diagnoses and systems diagnoses of both face-to-face and telerehabilitation physiotherapy examination. | 10 patients (9 males, 1 female; mean age = 38, SD = 13) recruited from a Physiotherapy Musculoskeletal and Sports Injury Clinic in Brisbane. |
| Lawford, 2017 <sup>49</sup> | Australia | Observational study | Questionnaire | Hip or knee osteoarthritis | A physical therapist-prescribed exercise program over the telephone and via video over the Internet (e.g. Skype, FaceTime). | (1) consumer perceptions and (2) willingness to use remotely delivered service models for physical therapist-prescribed exercise management of hip and/or knee osteoarthritis. | 330 patients (74 men and 253 women; mean age = 61.7; SD = 7.7) with hip or knee osteoarthritis. |
| Moffet, 2015 <sup>50</sup> | Canada | Randomized controlled trial (RCT) | Measurement of outcomes related to treatment efficacy to show the non-inferiority of telerehabilitation | Osteoarthritis | The rehabilitation intervention included 16 sessions of 45-60 minutes, delivered through an in-home telerehabilitation system with real-time 2-way videoconferencing, supervised by a trained physical therapist. The components of the intervention were an assessment before and after exercise, supervised exercises during a period of approximately 30 minutes, prescription of home exercises to perform on days without supervised sessions, and advice concerning pain control, walking aids, and the return to activities. | Primary outcome was the gain from baseline to the last follow-up in the Western Ontario and McMaster Universities Osteoarthritis Index<br>Secondary outcomes were changes from baseline to last follow-up in the Knee injury and Osteoarthritis Outcome Score. | 101 patients in the standard treatment group (56 males and 45 females; mean age = 67) and 104 patients in the telerehabilitation treatment group (44 males and 60 females; mean age = 65) recruited from the surgical waiting lists of orthopaedic surgeons from eight hospitals in Quebec, Canada. |
| Nelson, 2020 <sup>51</sup> | Australia | Randomized controlled trial (RCT) | Questionnaire | Hip osteoarthritis | Patients received a 6-week home exercise programme using the Wellpepper clinic, where the physiotherapist created exercise programmes for patients to follow on a tablet device. Participants were instructed to perform the home-based exercise program three times daily and self-monitor compliance using an exercise diary. During the 6 weeks, patients also received physiotherapy sessions via real-time videoconferencing using eHAB where analysis and advice regarding gait and their home exercise programme were undertaken. | Primary outcome was the quality of life subscale of the Hip disability and Osteoarthritis Outcome Score measured at six weeks post-operatively. Secondary outcomes included objective strength and balance outcomes, self-reported function and satisfaction outcomes, and home exercise program compliance. | 70 patients undergoing primary total hip replacement at the QEII Jubilee Hospital, Brisbane, Australia, of which 35 patients were in the intervention group (12 male and 23 female, mean age = 62) and 35 were in the control group (14 male and 21 female, mean age = 67). |
| Palacín-Marín, 2013 <sup>52</sup> | Spain | Mixed-methods study | Facilitators/barriers were not identified in the study. They were discussed generally in the discussion. | Chronic low back pain | Real-time online telerehabilitation assessments using Skype. In the online assessment, the examiner asked the participant to perform movements and functional tasks that enabled measurement of the outcome variables and offered real-time analysis of range of motion, quality of movement, and changes in symptoms. The online examiner made real-time corrections of leg and trunk positioning and technique quality and recorded patient responses on mechanosensitivity and the localization of symptoms. | Lumbar spine mobility, back muscle endurance, lumbar motor control, disability assessment, pain assessment, health-related quality of life, kinesiophobia, and telerehabilitation reliability and validity as compared to face-to-face assessment. | 15 patients (6 males and 9 females; mean age = 37) with chronic lower back pain from Gran Capitan primary care center of the Andalusian Health Service in Granada. |
| Pietrzak, 2013 <sup>53</sup> | Australia | Systematic review | Review of primary studies | Osteoarthritis | Internet-based physiotherapy interventions for adult patients with osteoarthritis. | This was a review of published literature investigating the effectiveness of community and home-based Internet interventions to self-manage, improve osteoarthritis-related health outcomes, and provide rehabilitation in osteoarthritis. | Adult patients with diagnosed osteoarthritis or osteoarthritis-related joint pain or those who underwent osteoarthritis-related joint replacement surgery from 5 studies included in the systematic review. |
| Russell, 2011 <sup>54</sup> | Australia | Randomized controlled trial (RCT) | Questionnaire | Degenerative arthritis of the knee | A six-week program of outpatient physical therapy delivered through an Internet-based telerehabilitation program. Participants in the telerehabilitation group received all rehabilitation through real-time interaction with a physical therapist across a low-bandwidth Internet-based telerehabilitation system. The intervention consisted of 1 treatment session with a duration of 45 minutes per week. A home exercise program that participants were encouraged to complete twice daily at home was also integrated in the treatment session. | The primary outcome was the Western Ontario and McMaster Universities Osteoarthritis Index questionnaire. Secondary outcomes were the Patient-Specific Functional Scale, the Spitzer Quality-of-Life Uniscale, the timed up-and-go test, and pain intensity rated on a visual analog scale. Physical measures were also recorded, including active and passive knee flexion and knee extension, quadriceps muscle strength assessed by knee extension lag during a straight-leg raise, girth measurements at the knee, and an assessment of gait with use of the Gait Assessment Rating Scale. | 34 patients in control group (mean age = 69.6; SD = 7.2) and 31 patients in telerehabilitation group (mean age = 66.2; SD = 8.4) who received a unicompartmental or unilateral total knee arthroplasty at a city hospital in Brisbane, Australia. |
| Russell, 2010a<sup>55</sup> | Australia | Mixed-methods study | Questionnaire and observation | Nonarticular lower-limb musculoskeletal disorders | Patients attended an online self-examination led by a physical therapist in real-time via a telerehabilitation system, during which patients performed functional and orthopedic tests with the assistance of the physical therapist remotely. | Primary outcomes were telerehabilitation validity (in comparison to face-to-face assessments) and reliability (both inter and intrarater).<br>Secondary outcome was patients' satisfaction with the telerehabilitation intervention. | 19 patients (5 males and 14 females; mean age = 26) with injuries to their lower limbs recruited from a Physical Therapy Musculoskeletal and Sports Injuries Clinic in Brisbane, Australia. |
| Russell, 2010b<sup>56</sup> | Australia | Observational study | Questionnaire and observation | Ankle pain and dysfunction | Patients received online assessments conducted by an online examiner, who instructed the participants to execute certain movements, functional tasks or modified self-orthopedic tests, then analyzed range of motion, quality of movement and provocation of pain or other symptoms in real time as well as having the option of reviewing a high-quality video clip (640 × 480 pixel resolution) of the task using the system's video capture and playback tool. | Outcomes were the criterion validity and reliability of conducting a remote musculoskeletal assessment of the ankle joint complex using telerehabilitation technologies compared with a face-to-face assessment. | 15 patients (5 males, 10 females; mean age = 24.5; SD = 10.8) with ankle pain who presented to a Musculoskeletal and Sports Injuries Clinic in Brisbane, Australia participated in the study. |
| Steele, 2012<sup>57</sup> | Australia | Randomized controlled trial (RCT) | Questionnaire and observation | Shoulder pain | Patients attended a single 1.5-hour session, including a patient interview, a face-to-face physical examination, and a remote physical examination. During the remote examination, patients received an online self-examination led by a physical therapist in real-time via a telerehabilitation system, during which patients performed functional and orthopedic tests with the assistance of the physical therapist in another room. | Primary outcomes were to evaluate use of a telerehabilitation system for valid and reliable diagnoses of shoulder disorders, establish validity and reliability of individual physical examination findings via telerehabilitation and to examine satisfaction of participants with the system for physiotherapy examination. | 22 patients with shoulder pain (16 males and 6 females; mean age = 30.7) recruited from the community of a large tertiary university hospital in Brisbane, Australia. |
| Taylor-Gjevre, 2017<sup>58</sup> | Canada | Randomized controlled trial (RCT) | Questionnaire | Rheumatoid arthritis | Videoconferencing participants received an in-person examination by an on-site | Outcomes included disease activity, quality of life, and patient satisfaction. | 85 patients (17 males and 68 females; mean age = 56.4; mean duration of RA = 13.9) |
|  |  |  |  |  | physical therapist who subsequently reported findings during the videoconferencing session with their urban-based rheumatologist. Patients then attended follow-up visits by videoconferencing with their urban-based rheumatologist and rural in-person physical therapist every 3 months over a 9-month period. Last, the rheumatologist examined patients in-person at 10 months after the initial visit. |  | with rheumatoid arthritis, of which 54 were randomized to the video-conferencing follow-up arm and 31 to the control arm. |
| Truter, 2014 <sup>59</sup> | Australia | Observational study | Questionnaire and comparison of physiotherapy assessments | Low back pain | Telerehabilitation assessments using the eHAB TR system (a computer-based videoconferencing system). | Primary outcomes were the concurrent validity of remote assessment of spinal posture, active movements of the lumbar spine, and the passive straight leg raise (SLR) test.<br>Secondary outcome was participant satisfaction with the TR assessment in a rural patient population (note that participants received both assessments at the same appointment, one after the other). | 26 patients (11 men, 15 women; mean age = 43) from a small town in regional Queensland, Australia. |
| Wong, 2005 <sup>60</sup> | China | Mixed-methods study | Questionnaire | Knee pain | The intervention comprised a weekly centre-based supervised session and home-based exercise programme, which was conducted once a week for a total of 12 sessions via videoconferencing, allowing real-time interaction between older subjects and the therapist. The contents of each session included reinforcement of home exercise, education on self-management of knee pain according to the guidelines of the American College of Rheumatology and peer group sharing. | Primary outcome was a self-reported standardized knee-specific questionnaire (the Western Ontario and McMaster Universities Osteoarthritis Index).<br>Secondary outcome measures include (1) range of motion of the knee, (2) quadriceps strength, (3) functional performance, (4) balance performance, (5) quality of life, (6) knowledge test, (7) exercise adherence, (8) level of acceptance of videoconferencing. | 20 older persons (2 males and 18 females; mean age = 75) with knee pain recruited from two community centres in Hong Kong. |

### Perceived Barriers

Out of the 23 publications, 20 (87%) reported perceived barriers of utilizing telerehabilitation for physical or occupational therapy.^38,40-50,52-59^ Overall, 41 different barriers were identified and grouped according to the CFIR domains and constructs (see table 2). Perceived barriers were categorized under all CFIR domains. Notably, the *Characteristics of Individuals* (*n* = 13 barriers, 32%), *Intervention Characteristics* (*n* = 11 barriers, 27%), and *Inner Setting* (*n* = 10 barriers, 24%) domains covered over 80% of the total barriers identified. In the *Characteristics of Individuals* domain, the most commonly identified construct was *Knowledge and Beliefs About the Intervention* (*n* = 8 studies, 35%), with privacy concerns being one of the most commonly reported barriers. In the *Intervention Characteristics* domain, the most commonly identified construct was *Design Quality and Packaging* (*n* = 11 studies, 48%) with specific barriers including technological issues, need for highly secure and efficient equipment, and patients finding the intervention program difficult to use or navigate. The most commonly identified construct in the *Inner Setting* domain was *Networks and Communication* (*n* = 9 studies, 39%), which included, for instance, impersonal connection and difficulty establishing rapport between patients and healthcare professionals. Clinicians reported specific barriers, including for example difficulty reading patient cues and body languages under the *Networks and Communication* in the *Inner Setting* domain, and examiner’s inexperience resulting in inaccurate diagnosis under the *Engaging* in the *Process* domain.

**Table 2.** Perceived barriers of telerehabilitation within the CFIR (N = 41)

| CFIR Domain | Construct | Barrier | Reference | Number of studies (%) |
| --- | --- | --- | --- | --- |
| Intervention Characteristics | Intervention Source | - | - | 0 |
|  | Evidence Strength and Quality | 1. patients' concerns about efficacy and comprehensiveness of telerehabilitation intervention | [40] | 3 (13.0%) |
|  |  | 2. lack of cost-benefit analysis | [44] |  |
|  |  | 3. efficacy of intervention for a sub-set of patients under certain circumstances | [50] |  |
|  | Relative Advantage | - | - | 0 |
|  | Adaptability | - | - | 0 |
|  | Trialability | - | - | 0 |
|  | Complexity | 1. time consuming intervention | [40] | 2 (8.7%) |
|  |  | 2. physiotherapists' concerns about ability to properly treat their patients | [40] |  |
|  |  | 3. complexities in setting up and using the equipment (ultrasonic device, camera) | [46] |  |
|  | Design Quality and Packaging | 1. technological issues/limitations (e.g., poor audio/visual quality) | [43, 45-48, 54-56, 58, 59] | 11 (47.8%) |
|  |  | 2. need for highly-secure and efficient equipment | [53] |  |
|  |  | 3. patients finding the intervention program difficult to use or navigate | [43] |  |
|  | Cost | 1. financial cost of the service | [49] | 2 (8.7%) |
|  |  | 2. high-cost of purchasing devices | [53] |  |
| Outer Setting | Patient Needs and Resources | 1. patients' other commitments (e.g., work, childcare) | [41] | 4 (17.4%) |
|  |  | 2. the potential need for more support for patients | [44] |  |
|  |  | 3. lack of time | [49] |  |
|  |  | 4. patients' preferences for more face-to-face sessions | [43] |  |
|  | Cosmopolitanism | - | - | 0 |
|  | Peer Pressure | - | - | 0 |
|  | External Policy and Incentives | - | - | 0 |
| Inner Setting | Structural Characteristics | - | - | 0 |
|  | Networks and Communications | 1. impersonal connection and difficulty establishing rapport between patients and healthcare professionals | [40, 42, 48, 55] | 9 (39.1%) |
|  |  | 2. lack of physical contact and non-verbal communication | [44, 49, 56] |  |
|  |  | 3. patients' poor communication skills during physical self-examinations | [55] |  |
|  |  | 4. difficulty reading patient cues and body languages | [42] |  |
|  |  | 5. dependence on local physiotherapist | [38] |  |
|  |  | 6. overreliance on information shared by patients rather than hands-on assessment | [45] |  |
|  | Culture | - | - | 0 |
|  | Implementation Climate | - | - | 0 |
|  | Readiness for Implementation | 1. waitlist management issues | [40] | 3 (13.0%) |
|  |  | 2. clinician availability issues | [40] |  |
|  |  | 3. the need to focus on videoconference aspect of the consultation (distraction) | [55] |  |
|  |  | 4. limited access to telerehabilitation technology in certain regions | [38] |  |
| **Characteristics of Individuals** | Knowledge and Beliefs About the Intervention | 1. privacy concerns/issues | [40, 42, 48, 52] | 8 (34.8%) |
|  |  | 2. concerns about the efficacy of videoconferencing | [49] |  |
|  |  | 3. insufficient knowledge on how to use videoconferencing services | [55] |  |
|  |  | 4. patients' preference of face-to-face assessment over remote assessment | [57] |  |
|  |  | 5. patients' and clinicians' preference for an initial in-person consultation | [45] |  |
|  | Self-Efficacy | 1. interference of other medical conditions | [41] | 4 (17.4%) |
|  |  | 2. patients' lack of confidence with videoconferencing or remote assessments | [49, 56] |  |
|  |  | 3. less compliance | [54] |  |
|  |  | 4. patients' difficulty in executing movements by the online examiner | [56] |  |
|  | Individual Stage of Change | - | - | 0 |
|  | Individual Identification with Organization | - | - | 0 |
|  | Other Personal Attributes | 1. patients' unwillingness to unrobe | [59] | 4 (17.4%) |
|  |  | 2. patients being reserved and less communicative in their conversations | [56] |  |
|  |  | 3. too much patient flexibility allowing for last-minute appointment cancellations | [45] |  |
|  |  | 4. patients' preference for travelling to their urban appointment locations | [58] |  |
| **Process** | Planning | - | - | 0 |
|  | Engaging | 1. examiners' inexperience resulting in inaccurate diagnoses | [57] | 2 (8.7%) |
|  |  | 2. high turnover of staffing | [38] |  |
|  | Executing | patients being not able to self-assess using current assessment tools | [55] | 1 (4.3%) |
|  | Reflecting and Evaluating | - | - | 0 |

### Perceived Facilitators

All 23 publications (100%) reported perceived facilitators in implementing telerehabilitation methods.^38-60^ In total, 59 facilitators were identified and categorized using the CFIR domains and constructs (see table 3), among which the *Intervention Characteristics* domain included the most reported facilitators (*n* = 26 facilitators, 44%), followed by the *Outer Setting* domain (*n* = 17 facilitators, 29%). In the *Intervention Characteristics* domain, *Design Quality and Packaging* (*n* = 10 studies, 43%) and *Cost* (*n* = 9 studies, 39%) were the two most commonly identified constructs, with example facilitators including audio and visual quality and financial savings, respectively. *Patient Needs and Resources* was the most commonly identified construct within the *Outer Setting* domain, within which time savings, the ability to access telerehabilitation from different locations, and convenience and accessibility of services were three of the most reported facilitators. Similar to perceived barriers, clinicians reported specific facilitators, for example, less staff travel costs under *Cost* in the *Intervention Characteristics* domain, and involving patients and getting their feedback under *Reflecting and Evaluating* in the *Process* domain.

**Table 3.** Perceived facilitators of telerehabilitation within the CFIR (N = 59)

| CFIR Domain | Construct | Facilitator | Reference | Number of studies (%) |
| --- | --- | --- | --- | --- |
| Intervention Characteristics | Intervention Source | - | - | 0 |
|  | Evidence Strength and Quality | 1. more service options | [50] | 3 (13.0%) |
|  |  | 2. sensitizing movements added to the intervention to aid diagnosis | [59] |  |
|  |  | 3. effectiveness of telerehabilitation intervention with a variety of devices | [53] |  |
|  | Relative Advantage | 1. flexibility to standard healthcare delivery | [40] | 5 (21.7%) |
|  |  | 2. videoconferencing builds better rapport between patients and healthcare providers, compared to telephone consultations | [40] |  |
|  |  | 3. maintenance of privacy | [39] |  |
|  |  | 4. less clinical atmosphere | [53] |  |
|  |  | 5. alternative solution to travelling | [57] |  |
|  |  | 6. less clinicians' unnecessary travels | [38] |  |
|  | Adaptability | tailored assessments for the purpose of telerehabilitation | [44] | 1 (4.3%) |
|  | Trialability | - | - | 0 |
|  | Complexity | 1. ease of use/easy-to-use equipment | [46, 49, 55, 59] | 8 (34.8%) |
|  |  | 2. readily accessible resources | [39, 43, 49, 51] |  |
|  |  | 3. offering devices to patients during the intervention | [53] |  |
|  |  | 4. user-friendly software | [51, 53] |  |
|  | Design Quality and Packaging | 1. audio and visual quality | [44, 45, 48, 51, 54, 56, 59] | 10 (43.4%) |
|  |  | 2. videoconferencing equipment supported secure encryption protocols | [53] |  |
|  |  | 3. online examiner sending a pre-made, high-quality video of the movement | [56] |  |
|  |  | 4. multi-site connection | [60] |  |
|  |  | 5. ability to demand remote, real-time feedback | [45, 47] |  |
|  |  | 6. ability to change volume to the participants' liking | [47] |  |
|  |  | 7. consistent and uninterrupted audio and video transmission | [47] |  |
|  | Cost | 1. financial savings (e.g., reduced costs associated with attending appointments) | [38-42, 44] | 9 (39.1%) |
|  |  | 2. cost-effectiveness | [38, 50] |  |
|  |  | 3. affordability of videoconferencing equipment | [53] |  |
|  |  | 4. free software | [52] |  |
|  |  | 5. less staff travel costs | [38] |  |
| **Outer Setting** | Patient Needs and Resources | 1. less patients' unnecessary travels | [38, 40, 58] | 16 (69.6%) |
|  |  | 2. time savings | [41, 49, 50, 55, 58] |  |
|  |  | 3. ability to access telerehabilitation from different locations (e.g., home, work) | [41, 45] |  |
|  |  | 4. faster access to recommended healthcare services | [41] |  |
|  |  | 5. patient travel savings | [38, 42, 44] |  |
|  |  | 6. detailed description to educate patients on telerehabilitation | [44] |  |
|  |  | 7. including in-person visits with telehabilitation that may be beneficial for patient care | [44] |  |
|  |  | 8. assured privacy | [49] |  |
|  |  | 9. friend or untrained nonclinical assistant accompanying the participant | [59] |  |
|  |  | 10. presence of a local physiotherapist at the preliminary consultation | [38] |  |
|  |  | 11. shorter travelling distance | [53] |  |
|  |  | 12. transport access | [53] |  |
|  |  | 13. convenience and accessibility of services | [38, 45, 46, 48, 51] |  |
|  |  | 14. sufficient time to answer patients' questions | [42] |  |
|  |  | 15. removal of distractions | [47] |  |
|  |  | 16. patients receiving technological assistance including trouble-shooting difficulties | [45] |  |
|  |  | 17. saved waiting time for clinical appointments | [38] |  |
|  | Cosmopolitanism | - | - | 0 |
|  | Peer Pressure | - | - | 0 |
|  | External Policy and Incentives | - | - | 0 |
| **Inner Setting** | Structural Characteristics | - | - | 0 |
|  | Networks and Communications | 1. improvement in communication between healthcare professionals | [40] | 4 (17.4%) |
|  |  | 2. improved and sustained connection with healthcare practitioners | [41] |  |
|  |  | 3. development of a bond with the therapist | [39] |  |
|  |  | 4. online examiner providing real-time feedback | [56] |  |
|  | Culture | - | - | 0 |
|  | Implementation Climate | patients being comfortable and relaxed in home environment | [45] | 1 (4.3%) |
|  | Readiness for Implementation | 1. educational opportunity and development of internal focus of control | [54] | 2 (8.7%) |
|  |  | 2. detailed patient interview information and accurate physical objective observations | [56] |  |
| **Characteristics of Individuals** | Knowledge and Beliefs About the Intervention | 1. perceived benefit of intervention | [54] | 5 (21.7%) |
|  |  | 2. information can be easily understood | [42] |  |
|  |  | 3. high patient satisfaction towards telerehabilitation | [43, 51, 58] |  |
|  |  | 4. high compliance with telerehabilitation intervention | [51] |  |
|  | Self-Efficacy | patients being comfortable with telecommunication technologies | [43] | 1 (4.3%) |
|  | Individual Stage of Change | - | - | 0 |
|  | Individual Identification with Organization | - | - | 0 |
|  | Other Personal Attributes | - | - | 0 |
| **Process** | Planning | - | - | 0 |
|  | Engaging | 1. interprofessional team members to facilitate the telerehabilitation | [44] | 3 (13.0%) |
|  |  | 2. availability of a trained assistant | [56] |  |
|  |  | 3. trained therapists | [55] |  |
|  | Executing | - | - | 0 |
|  | Reflecting and Evaluating | involving patients and getting their feedback | [44] | 1 (4.3%) |

## Discussion

The results of this scoping review suggest that telerehabilitation can be used effectively to deliver physical or occupational therapy interventions and assessments for musculoskeletal diseases when important barriers, such as audio and visual quality, are considered. Many patients appreciate the convenience and flexibility of telerehabilitation when it reduces the need to travel, leading to time and financial savings. However, key barriers in the implementation and adoption of telerehabilitation interventions were identified, among which the most mentioned ones were technological issues (e.g. poor audio or visual quality), and concerns about confidentiality and privacy, as well as a less personal connection and limited confidence in using technology. These barriers are in line with findings from other studies in the literature.^61-64^

Several other studies across settings have been conducted to examine the barriers and facilitators of delivering telerehabilitation intervention programs to patients. These studies have shown that the intention to use technology and the uptake and implementation of technological innovations in practice are mainly predicted by factors such as perceived usefulness, perceived ease of use, user experience, and level of technology confidence.^65-67^ In the development and implementation of telerehabilitation intervention programs, addressing these factors when patients are invited to try novel online interventions may lead to higher user satisfaction and improve uptake and adherence. Our results suggest that the quality and user-friendliness of the intervention are key factors to be considered, and that sufficient resources and training need to be provided to support both patients and clinicians in using telerehabilitation applications efficiently.

There are several limitations that should be considered when interpreting the results of this review. First, by definition, research questions addressed by scoping reviews are often exploratory in nature, identifying all relevant literature regardless of study design. Most of the included studies were relatively small studies that were done in different musculoskeletal diseases and used different methodologies; some used questionnaires or interviews to obtain data, and some reflected the experience of the authors without collecting data systematically.

Second, although we had data extraction tools and two different investigators extracting data independently from publications, extracting accurate and complete data remained a challenge. There are a number of reasons for this, including that some articles reported methods or results that were incomplete or unclear. In addition, challenges occurred while investigators were identifying the most accurate CFIR domains and constructs for certain barriers and facilitators when both constructs appeared to be reasonable. For instance, “financial savings” was categorized under *Cost* by one investigator, but under *Patient Needs and Resources* by the other. To overcome this, coders discussed and consulted with the third investigator to make the final decision. Finally, we had to limit our scoping review to synchronous videoconferencing only due to the high number and variability of applications that used different types of technologies. Indeed, a systematic review identified 51 unique definitions of e-health which showed a wide range of themes, but there was no clear consensus about the meaning of the term e-health or the technologies and intervention processes often described as e-health interventions.^68^ One of these definitions, that we had originally started to use in our project, appeared to be too broad for our purposes. We therefore refined our definition and did several rounds of pilot testing to ensure it would adequately capture the studies that would inform the development of a physical or occupational therapy intervention involving direct contact between patients and health care providers and exclude others.

## Conclusion

In sum, telerehabilitation interventions delivered via videoconferencing with a healthcare professional can be an effective way to deliver patient-centered physical or occupational therapy telerehabilitation interventions for people with SSc and other common or rare musculoskeletal conditions. However, the findings of this scoping review present an overview of some important barriers that need to be addressed in order to enhance the development and implementation of telerehabilitation programs for patients, including people with SSc. Strategies to combat barriers to using telerehabilitation may include using a stable, high-quality videoconferencing platform, enhancing self-efficacy to using videoconferencing amongst patients and health care providers, and addressing concerns related to privacy. During the current COVID-19 pandemic, using telerehabilitation has become an increasingly important strategy to deliver rehabilitation interventions and assessments for at-risk populations, and the trend will likely continue to grow during and after the pandemic. Therefore, the present study may also provide useful insight into the successful development and delivery of physical or occupational telerehabilitation interventions when in-person appointments are not easily conducted.

## Data Availability

No data were produced for this scoping review (use of published data only)

## Acknowledgements

We thank Naz Torabi, Information Specialist at Unity Health Toronto, for peer review of the MEDLINE search strategy.

## Declaration of Interest

The authors declare that they have no conflict of interest.

## Funding

This work was supported by an Edith Strauss Project Grant. Ms. Carboni-Jiménez was supported by a McGill University Faculty of Medicine Solvay Fellowship and by a McGill University Delta Upsilon Scholarship. Dr. Thombs was supported by a Tier 1 Canada Research Chair.

## Appendix 1. Search terms

**(musculoskeletal or bone or osteochondritis or cartilage or ligaments or collagen or fasciitis or foot deformit* or metatarsal deformit* or hand deformit* or (jaw adj1 (disease* or disorder*)) or arthropath* or arthroses or arthrosis or (joint* adj1 (disease* or disorder* or deformit*)) or (muscle adj1 (disease* or disorder*)) or (muscular adj1 (disease* or disorder*)) or myopath* or rheumatic or rheumatism or arthritis or fibromyalgia or polymyalgia or scleroderma or systemic sclerosis or connective tissue or Osteoarthritis)**

**(musculoskeletal or bone or osteochondritis or cartilage or ligaments or collagen or fasciitis or foot deformit* or metatarsal deformit* or hand deformit* or (jaw N1 (disease* or disorder*)) or arthropath* or arthroses or arthrosis or (joint* N1 (disease* or disorder* or deformit*)) or (muscle adj1 (disease* or disorder*)) or (muscular adj1 (disease* or disorder*)) or myopath* or rheumatic or rheumatism or arthritis or fibromyalgia or polymyalgia or scleroderma or systemic sclerosis or connective tissue or Osteoarthritis)**

((mobile adj1 (health or rehabilitation)) or tele health or telehealth or telemedicine or e-health or ehealth or m-health or mhealth or mobile based or (virtual adj1 (medicine or rehabilitation)) or (remote adj1 (medicine or rehabilitation or consultation)) or videoconferenc* or video conferenc* or tele conferenc* or teleconferenc* or tele education)

((mobile N1 (health or rehabilitation)) or tele health or telehealth or telemedicine or e-health or ehealth or m-health or mhealth or mobile based or (virtual N1 (medicine or rehabilitation)) or (remote N1 (medicine or rehabilitation or consultation)) or videoconferenc* or video conferenc* or tele conferenc* or teleconferenc* or tele education)

## APPENDIX 2. Coding Manual

### No: not a primary report or review on telerehabilitation (telehealth or e-health) methodsa

If the article does not report on telerehabilitation methods, it will be excluded. To be eligible reports on interventions must evaluate barriers and facilitators of using telerehabilitation methods. Telerehabilitation interventions, sometimes describe as telehealth or e-health interventions, include delivery of care from a distance to support, educate, inform and connect health care professionals and the people they serve through the use of information communications technologies. Eligible intervention delivery methods include online video conferencing, online interventions, mobile phone apps (mHealth), and the use of remotely monitored rehabilitation devices to deliver online rehabilitation interventions. For this review, telerehabilitation methods must involve a component of direct clinician and patient interaction via telerehabilitation methods and may or may not also include self-guided online material. Eligible activities include instruction, intervention, or assessment delivered via telerehabilitation methods. If telerehabilitation methods are not used for these purposes, the article will be excluded. Interventions that involve telephone interactions only will be excluded. Methods that only involve clinician-to-clinician consultation or training are excluded. Study protocols, instructional documents, conference abstracts, editorials and letters will be excluded.

### No: not an intervention for individuals with a musculoskeletal disease

If the article is not about a telerehabilitation intervention intended to help people with musculoskeletal diseases then it is excluded. Musculoskeletal diseases are diseases that affect the human body’s movement or musculoskeletal system. Only articles about diseases of the musculoskeletal system and connective tissue, defined per the International Statistical Classification of Diseases and Related Health Problems, 10th revision (ICD-10; M00-M99) will be included. Studies including a mixed population of people with and without a musculoskeletal disease are excluded unless results for people with a musculoskeletal disease and people without a musculoskeletal disease are reported separately.

### No: not a physical or occupational therapy intervention or assessment

If the article is not examining a physical or occupational therapy intervention or a physical or occupational therapy assessment to develop a physical or occupational therapy program, then it is excluded.

<u>Eligible occupational therapy interventions</u> include interventions described as an occupational therapy intervention and interventions delivered by an occupational therapist or under the supervision of an occupational therapist. Occupational therapy interventions focus on activities of daily living (ADL), which may include eating and drinking, functional mobility, going to the toilet, dressing, carrying out personal hygiene and grooming activities, as well as instrumental activities of daily living (IADL), such as education, work, play, leisure, and social participation). <u>Eligible occupational therapy assessments</u> are conducted by an occupational therapist for the purpose of designing, adapting, or evaluating an occupational therapy intervention. Occupational therapy interventions, procedures and assessments include:

1. Occupational therapy assessment to determine abilities and impairments, which may include history-taking, physical tests and measures, or standardized assessments;
2. Treatments focused on remediating impaired capacities or abilities, such as task-orientated approaches or activity-based interventions;
3. The use of adaptive (compensatory) techniques as alternative ways to effectively complete ADLs;
4. The use of assistive technology, in which the patient has the use of any item, piece of equipment, or product system to increase, maintain, or improve their functional capacity; and
5. Environmental adaptations in which the participant’s physical environment is modified with ramps, electric hoists, stair lifts, handrails, level access, or a wet-room wheelchair accessible shower in order to restore or enable self-reliance, privacy, confidence, or dignity.

<u>Eligible physical therapy interventions</u> include interventions described as a physical therapy intervention and interventions delivered by a physical therapist or under the supervision of a physical therapist. <u>Eligible physical therapy assessments</u> are conducted by a physical therapist for the purpose of designing, adapting, or evaluating a physical therapy intervention. Physical therapy interventions and assessments include:

1. Physical therapy assessments to determine abilities and impairments, which may include history-taking, physical tests and measures and/or standardized assessments;
2. The use of therapeutic exercise such as aerobic capacity/endurance conditioning or reconditioning; balance, coordination, and agility training; body mechanics and postural stabilization; flexibility exercises; gait and locomotion training; neuromotor development training; relaxation; strength, power, and endurance training;
3. Functional training in self-care and home management such as activities of daily living; barrier accommodations or modifications; device and equipment use and training; functional training programs, injury prevention or reduction; and
4. Functional training in work (job/school/play), community, and leisure integration or reintegration such as barrier accommodations or modifications; device and equipment use and training; functional training programs; injury prevention or reduction; manual therapy techniques including: passive range of motion; massage; mobilization/manipulation.

Multifaceted programs that include a substantial component of physical or occupational therapy that would be eligible are included.

**No: not a study or report that discusses barriers or facilitators for using telerehabilitation (telehealth or e-health) in clinician-to-patient interactions**. If the article does not describe barriers or facilitators to delivering direct, synchronous clinician-to-patient interaction via online videoconferencing, then it is excluded. Articles mentioning only regulatory or policy barriers are excluded.

**Yes: study eligible to be included in scoping review**.

## Notes

### Competing Interest Statement

The authors have declared no competing interest.

